# Teaching Ultrasound Early: Outcomes from a Student-Led POCUS Elective Course

**DOI:** 10.64898/2026.01.16.26344280

**Authors:** Ikaasa Suri, Nicole Parkas, Elise Solazzo, Ryan Yu, Neal Moehrle

**Author notes:** **Corresponding Author:** Ikaasa Suri, B.S., Mailing Address: Icahn School of Medicine at Mount Sinai, Department of Emergency Medicine, 1468 Madison Avenue, New York, NY 10029 New York, NY, 10029 USA.

## Abstract

**Background:** Point-of-care ultrasound (POCUS) utilization across specialties continues to grow, making it a valuable skill for medical students. Early exposure to ultrasound may enhance students’ clinical reasoning, anatomical understanding, and integration of POCUS into the physical exam. This study evaluates the impact of a student-organized, physician-taught POCUS elective course on pre-clinical medical students’ competence in foundational ultrasound skills.

**Methods:** Fifteen students were enrolled in the course. Students attended four weekly 90-minute sessions focused on a unique organ system. Students took a 19-question test before and after the course to assess overall learning. Five-question quizzes were conducted before and after each session to evaluate immediate learning. Fisher’s exact test was used to compare correct vs. incorrect quiz answers before and after each session.

**Results:** Overall knowledge of POCUS, determined by the 19-question quiz, improved from 61.5% to 76.8% (p = 0.01). FAST and cardiac ultrasound quiz scores improved from 54.2% to 91.4% (p < 0.01) and 57.7% to 85.7% (p < 0.01), respectively. The pre- and post-quiz score for the abdominal ultrasound exam remained the same at 80.0% (p = 1). The gynecologic ultrasound exam score improved from 45.0% to 66.7% (p = 0.3).

**Conclusions:** This elective course significantly increased pre-clinical medical students’ knowledge of ultrasound. No statistically significant difference was noted for the abdominal or gynecologic sessions.

## 1. Introduction

The utilization of point-of-care ultrasound (POCUS) has become virtually ubiquitous among medical specialties today, making it an extremely valuable skill for medical students to learn. Early exposure to ultrasound in undergraduate medical education (UME) may aid in clinical reasoning, anatomical understanding, and physical exam integration as students transition from the classroom to clinical settings [1,2]. Additionally, there has been a movement among residency programs to incorporate ultrasound education into their curricula [3-6]. In response, many UME institutions have expanded ultrasound exposure within their curricula to prepare students [7-9].

The benefits of POCUS training in UME extend far beyond preparation for residency. A six-year study done at Ohio State University assessed long-term benefits from UME ultrasound exposure in medical students who underwent a fourth-year ultrasound elective compared to classmates who had not. The study found that students who participated in the elective were more likely to use ultrasound in clinical decision making, to apply it in emergency settings, and to seek additional training during residency [10]. Developing ultrasound skills and confidence early in medical training promotes greater use in future practice, which can ultimately improve diagnostic accuracy and patient care.

The current state of POCUS education in UME is well documented in the literature. A national survey of allopathic medical schools in the United States showed the number of medical schools incorporating POCUS in their curriculum increased from 51 in 2012 to 84 in 2025. Additionally, the number of schools implementing a longitudinal ultrasound curriculum rose from 10 in 2021 to 13 in 2025. Nevertheless, the number of schools evaluating students’ understanding of POCUS did not increase from 5 years ago. Even with increased incorporation of ultrasound education, there still remains a lack of research on POCUS incorporation and standardized guidelines for medical schools to follow [7].

It has been reported that the most prominent barriers to POCUS incorporation in UME curricula are lack of trained faculty, lack of time in current curricula, and lack of equipment [11]. However, studies have shown successful implementation of ultrasound education with limited faculty, such as utilizing student leadership and organization as a means to incorporate education in a resource-limited setting [12]. To explore the role of students in facilitating ultrasound education under resource-limited conditions, we developed a four-week, student-organized, physician-taught ultrasound elective course for first- and second-year medical students. The objective of this study was to evaluate how student leadership can support the acquisition of foundational ultrasound skills in a resource-limited setting.

## 2. Methods

### 2.1 Study Population

Fifteen novice first- and second-year medical students were enrolled in our elective course, organized over four weeks. None of the students had any prior exposure to ultrasound, but all had completed their anatomy module before enrolling. This study was approved by the Institutional Review Board at Mount Sinai Hospital.

### 2.2 Course Content

Students in this course first received a didactic general introduction to ultrasound that covered ultrasound terminology, probe/machine basics, and knobology. Successive weeks focused on a specific POCUS application: the Focused Assessment with Sonography for Trauma (FAST) exam, echocardiography, the abdominal (biliary, renal, and abdominal aorta) exam, and OB/GYN exams. Each session consisted of a 30-minute didactic lecture and an hour-long hands-on skills session provided by either faculty or emergency medicine residents credentialed in each particular modality, as determined by the American College of Emergency Physicians (ACEP) guidelines. Students were divided into two groups during the hands-on portion to optimize individual learning.

### 2.3 Questionnaires and Statistics

A 19-question multiple-choice quiz was administered at the beginning and at the conclusion of the course. Additionally, 5-question multiple-choice quizzes were administered at the beginning and end of each system-specific session to assess students’ understanding of focused ultrasound skills. These questions are listed in **Appendix A**. Fisher’s exact test was used to compare the number of correct and incorrect assessment answers before and after each class, and upon course completion.

## 3. Results

While fifteen pre-clinical medical students were enrolled in the four-week, student-organized, physician-taught POCUS elective, attendance and questionnaire response rates varied per session. Overall knowledge, assessed by a 19-item quiz administered before and after the course, improved from 61.5% to 76.8% (p=0.01). Session-specific quizzes showed significant gains for the FAST exam (54.2% to 91.4%, p<0.01) and cardiac ultrasound (57.7% to 85.7%, p<0.01). Abdominal ultrasound scores were unchanged (80.0% to 80.0%, p=1.0). Gynecologic ultrasound scores increased from 45.0% to 66.7%, which did not reach statistical significance (p=0.30). Pre- and post-session results are shown in **Figure 1**.

**Figure 1.**
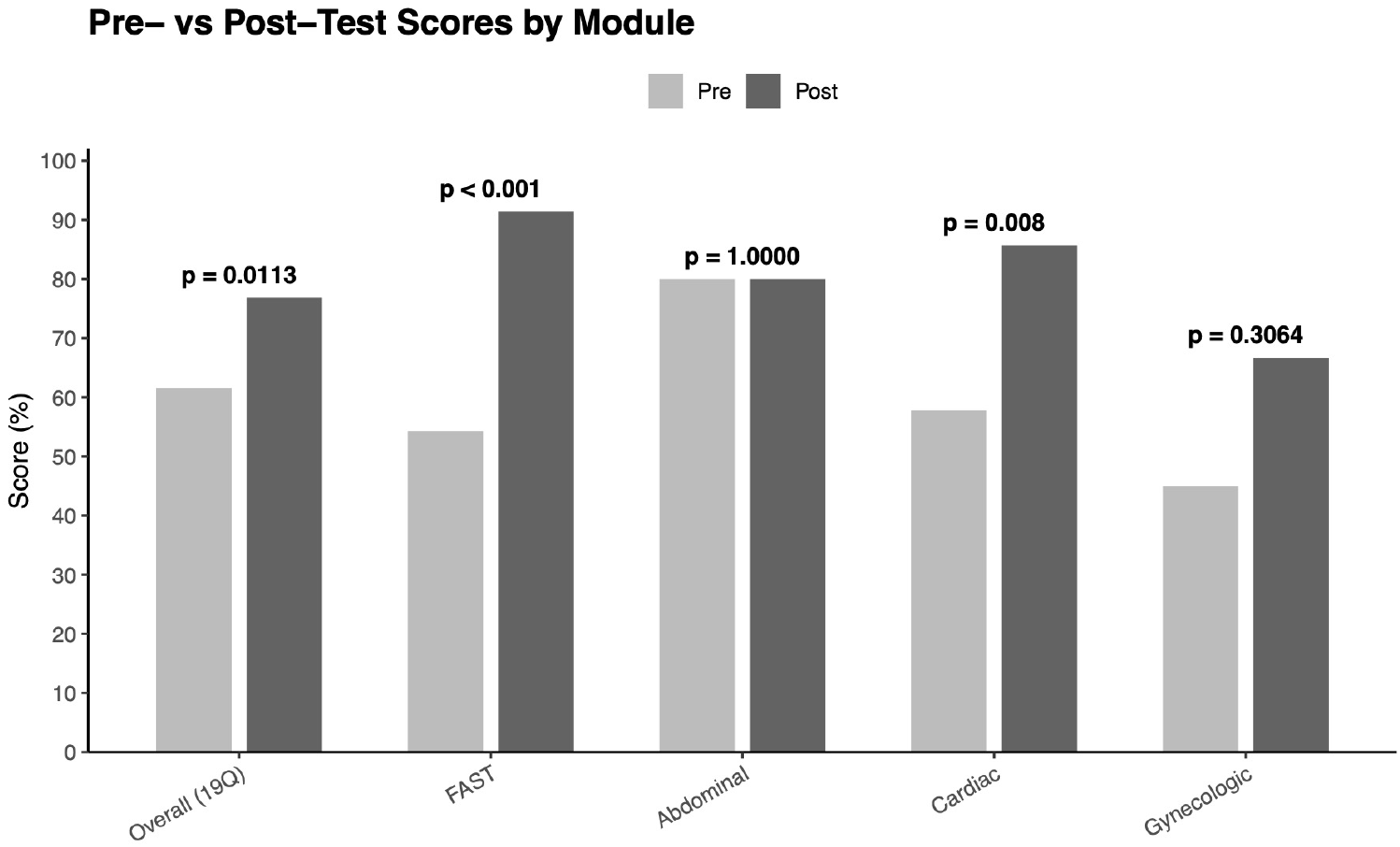
Average pre- and post-test scores by course module. P-values were calculated from Fischer’s exact test comparing distribution of correct and incorrect answers pre vs. post-instruction. The overall, 19-question pre- and post-class survey, as well as all subject data analyses utilized data from all available students. Sample sizes: Overall (19Q) Pre n = 10, Post n = 5; FAST Pre n = 7, Post n = 7; Abdominal Pre n = 3, Post n = 3; Cardiac Pre n = 9, Post n = 5; Gynecologic Pre n = 4, Post n = 3.

## 4. Discussion

In this pilot of a student-organized and physician-taught POCUS elective, participants demonstrated meaningful gains in overall foundational ultrasound knowledge and in two core applications: the FAST exam and echocardiography. These findings align with the broader literature which shows that targeted, early POCUS instruction improves medical students’ knowledge, confidence, and readiness for clinical application. Prior work has reported that integrating POCUS into undergraduate medical education enhances anatomic understanding and clinical reasoning and can translate to better performance on knowledge tests and structured skills assessments, such as direct clinical observations and OSCEs [8,13].

The most improvement amongst students was illustrated in the FAST and cardiac modules, which is consistent with studies demonstrating substantial short-term knowledge and performance gains after focused FAST training and cardiovascular POCUS teaching [14-15]. Our study specifically used live modules; however, in some settings, students trained on the FAST exam achieve comparable written and OSCE scores whether instruction uses live models or simulation, supporting flexibility in delivery methods [16]. Additionally, recent studies suggest that prior ultrasound exposure potentiates gains during subsequent FAST training, reinforcing the value of introducing POCUS early in the curriculum [17].

The lack of detectable change in the abdominal ultrasound module likely reflects a ceiling effect as evident by high baseline scores, while the non-significant gynecologic gains are plausibly attributable to limited sample size and variable attendance—both common challenges in pilot electives. Similar implementation studies emphasize that feasibility constraints (e.g. protected time, equipment, funding, and availability of trained faculty) shape outcomes as much as pedagogy [18-20]. Low-resource and hybrid models–brief didactics plus hands-on scanning, near-peer components, and tele-enabled or asynchronous elements–have been successfully deployed and may help scale offerings without sacrificing learning [16-17].

Our student-organized structure is supported by literature showing that student-led or near-peer POCUS teaching can achieve learning outcomes comparable to faculty-only instruction while expanding capacity and leadership development [21]. A randomized comparison of near-peer versus faculty instruction found similar effectiveness across multiple applications, and a dedicated student-led curriculum reported significant pre- and post-knowledge gains across sessions [12].

For medical schools seeking to introduce POCUS early, a concise, focused elective can yield measurable gains in high-value applications such as the FAST exam and basic echocardiography. Embedding structured practice, brief assessments, and opportunities for deliberate scanning likely augments retention and transfer to clinical settings; contemporary studies of other focused protocols show knowledge and simulator improvements but caution that clinical proficiency may require longitudinal reinforcement [22].

### 4.1 Limitations

This single-institution pilot had a small cohort and relied on short knowledge quizzes rather than assessment of image acquisition. Additionally, our elective course was offered in conjunction with the students’ required medical school coursework, which likely impacted session attendance. In turn, there was limited participant completion of the pre- and post-course tests. Fisher’s exact tests were appropriate for small samples but limited covariate adjustment. Future work should incorporate performance-based endpoints (e.g., validated ultrasound OSCEs), track knowledge and skill retention, and evaluate patient-proximal outcomes during clerkships.

### 4.2 Conclusion

A physician-taught, student-organized POCUS elective produced significant knowledge gains overall and in the FAST and cardiac modules among pre-clinical students. Results support integrating POCUS early in undergraduate medical education using scalable delivery models—potentially including near-peer elements—to build foundational skills that can be reinforced longitudinally.

## Data Availability

All data produced in the present work are contained in the manuscript.

## Ethics Statement

All participants enrolled in this study were consented according to the IRB approved by Mount Sinai.

## Disclosures

Ikaasa Suri is a stockholder at Illuminant Surgical, Inc. However, Illuminant Surgical is not impacted by nor has it funded this research in any manner.

## Notes

### Funding Statement

This study did not receive any funding.

